# Rare Coding Variants Reveal Distinct Genetic Architectures Across Multidimensional Sleep Phenotypes

**DOI:** 10.64898/2026.06.16.26355625

**Authors:** Yingzhe Zhang, Wenhan Lu, Lovemore Kunorozva, Samuel Jones, Matthew Maher, Jesse Valliere, Andrew R. Wood, Michael N. Weedon, Justin Tubbs, Konrad J. Karczewski, Tian Ge, Henning Tiemeier, Jacqueline Lane, Richa Saxena, Hanna M Ollila, Chia-Yen Chen

## Abstract

Sleep and circadian traits have been widely studied using common variants, but the contribution of rare coding variation remains unclear. We analyzed rare coding variants in 397,065 whole-exome sequenced UK Biobank participants across 36 sleep phenotypes from self-report, diagnoses, sleep medication use and accelerometry, and meta-analyzed results with 171,536 whole-genome sequenced All of Us participants of diverse ancestries, with replication in the Mass General Brigham Biobank (N = 31,275). We identified 260 genes associated with sleep phenotypes, including novel associations with sleep medication use in 29 genes and 24 out of 29 have not previously been reported with any sleep phenotypes. We observed modest but significant rare variant heritability and strong genetic correlations between sleep medication use, insomnia and fatigue. Temporal gene expression trajectory analyses indicate that genes associated with self-reported sleep traits show constant high prenatal expression, whereas genes linked to sleep medication phenotypes exhibit peak expression in the late prenatal period. These findings highlight distinct biological mechanisms captured by different measurement sources of sleep phenotypes and reveal rare-variant-informed targets for therapeutic discovery.

## Main

Genome-wide association studies (GWAS) have been widely conducted to uncover the genetic architecture of common variants underlying sleep and circadian phenotypes. Large-scale GWAS have revealed numerous loci associated with sleep duration, chronotype, insomnia, and circadian rhythm, implicating pathways linked to synaptic signaling, neurotransmitter regulation, and circadian rhythm genes.^1–3^ In addition to these population-based findings, high-impact Mendelian mutations identified in family studies have provided mechanistic insights.

Despite such progress, the contribution of rare coding variations to a broader set of sleep-related phenotypes at the population level remains only partially characterized. In particular, its impact on severe clinical sleep disorders and sleep medication use remains unclear. While conventional GWAS approaches are restricted to common variants, sequencing-based studies are beginning to systematically identify rare exonic variants associated with complex traits through burden association testing within large cohorts.^4,5^ However, only one previous exome sequencing study reported significant findings in self-reported sleep phenotypes, leaving a wide spectrum of sleep and circadian phenotypes uninvestigated for rare variant associations.^6^ This limitation creates a critical gap in our understanding of how rare, and potentially deleterious, genetic variation shapes not only the continuum of phenotypic variation in traits such as sleep duration, timing, and stability but also extreme sleep disorders and medication use. A more comprehensive phenotyping approach is essential for capturing the full complexity of sleep behaviors and disorders. Many sleep behavior phenotypes show moderate to strong genetic correlations among common variants, with shared loci contributing to multiple traits across different assessment methods.^7–9^ Using a broad set of sleep measures helps disentangle trait-specific and pleiotropic genetic effects, clarifying how rare coding variants impact diverse sleep characteristics. More specifically, leveraging electronic health records (EHR) linked to large-scale biobanks enables the systematic investigation of severe sleep disorders and medication use, providing a powerful framework to delineate the contribution of rare coding variation to clinically ascertained sleep phenotypes.

Here we report an exome-wide association study (ExWAS) on a wide range of sleep phenotypes, including four different measurement modalities: self-reported phenotypes, sleep disorder diagnoses, sleep medication and accelerometer-based sleep behavior measures. By integrating population-based sequencing data with functional annotation, pathway enrichment and developmental gene expression trajectory analyses across sleep phenotypes from four different measurement sources, we systematically characterize the rare coding genetic architecture of sleep. This systematic identification and comparison of rare coding across different measures deepens our understanding of the genetic basis of sleep and highlights potential targets for precision medicine and therapeutic development.

## Results

### Overview of Study Design

To identify sleep genes across a broad spectrum of sleep phenotypes, we performed an exome-wide association study (ExWAS) to investigate the rare-coding genetic basis of 36 sleep-related phenotypes in the UK Biobank (UKB; N = 397,065). We identified sleep phenotypes from four types of measures: self-reported measures, clinically diagnosed disorders, medication use, and accelerometer-derived phenotypes (Methods). These phenotypes span a broad spectrum of sleep behaviors, from chronotype and napping to diagnoses such as insomnia, restless legs syndrome, and hypersomnia, as well as the use of common sleep medications (e.g., melatonin, zopiclone) and objective assessments of sleep quality (**Supplementary Table 1**). Notably, this represents the first exome-wide effort to systematically investigate the rare-coding genetic basis of sleep-related medication phenotypes. To enhance discovery power, we performed multi-ancestry meta-analyses with the All of Us Research Program (AoU, N = 171,536). We also further performed replication in exome sequencing data from the Mass General Brigham Biobank (MGBB, N = 31,275). Then, we characterized the rare-coding genetic architecture of sleep phenotypes by estimating rare variant burden heritability and rare variant burden genetic correlations in UKB. Finally, we conducted pathway enrichment network analyses and temporal brain-based gene expression trajectories across the lifespan for each measurement type to provide biological context for our genetic findings. An overview of the study design is presented in **Figure 1**.

**Figure 1.**
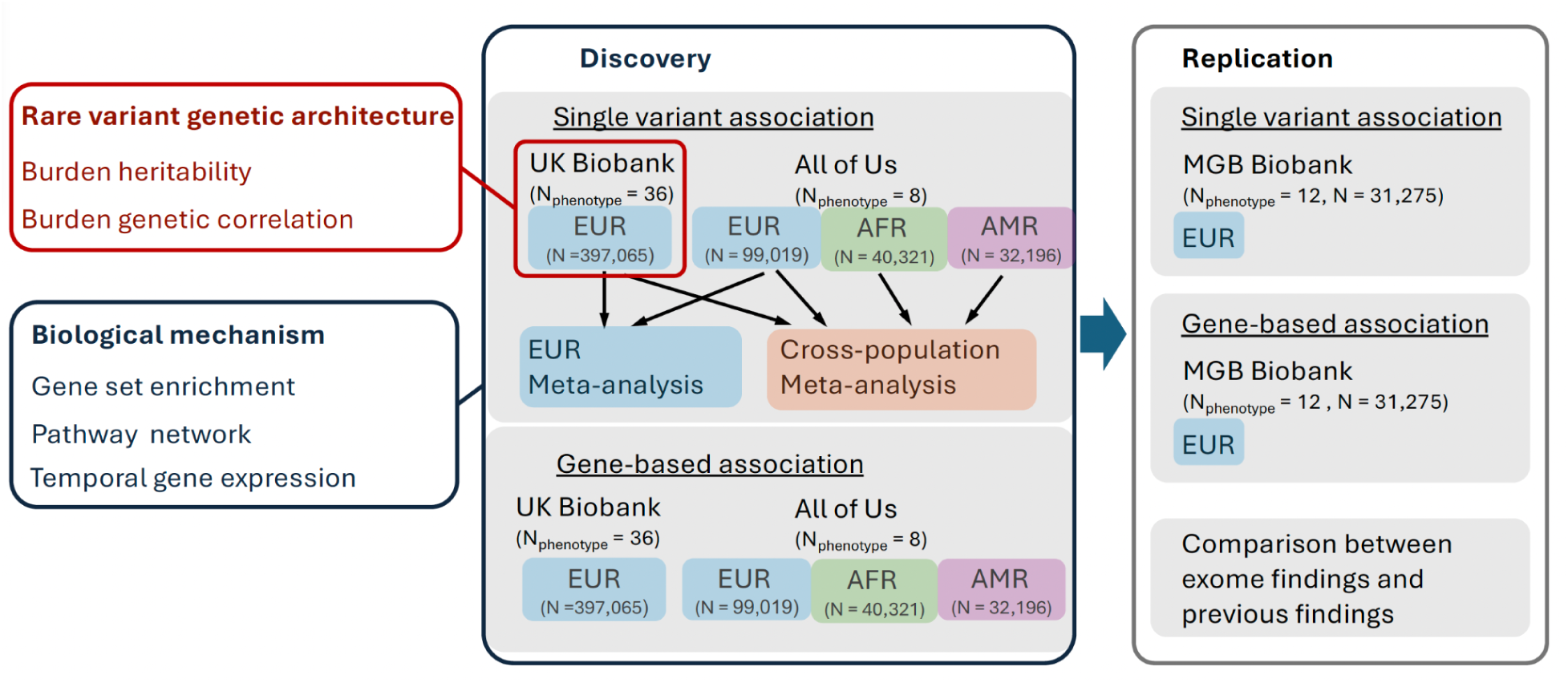
Analysis workflow for sleep exome associations in the UK Biobank, the All of Us Research Program (AoU), and the Massachusetts General Brigham (MGB) Biobank. First, both single-variant and gene-based association analyses were conducted for all 36 phenotypes in the UK Biobank. Next, single-variant association analyses for 8 sleep disorders were performed in AoU for European, African, and Admixed American ancestry groups (indicated in different colors) and meta-analyzed with the corresponding phenotypes in the UK Biobank. To replicate the results, we conducted both single-variant and gene-based association analyses in MGBB for the significant findings from the discovery stage. We also conducted follow-up analyses, including rare variant burden heritability analyses and biological mechanism analyses.

### Exome-wide Association Studies for 36 Sleep-Related Traits

We performed single variant association tests for 36 sleep phenotypes using whole-exome sequencing data from UKB participants of European ancestry (N = 397,065). Exome sequencing data in the UKB was annotated across three variant functional categories: protein-truncating variants (PTVs), damaging missense variants (classified as deleterious with CADD score > 20), and synonymous variants, with association tests performed for PTVs and damaging missense rare variants. Similarly, we also performed single variant association tests for 8 sleep disorder diagnoses using whole-genome sequencing data from AoU. In AoU, exome variants were annotated for 99,019 individuals of European ancestry, 40,321 of African ancestry, and 32,196 of Admixed American ancestry in three categories: PTVs, missenseLC (combined missense variants and low confidence PTVs), and synonymous variants. We also conducted inverse-variance weighted meta-analyses of single variant associations separately for UKB-AoU European ancestry samples and UKB–AoU cross-ancestry samples. Rare coding variants were defined as those with minor allele frequency (MAF) less than 0.01 in each cohort. To ensure robust statistical inference, we filtered the single variant association tests and only reported genome-wide significant findings for variants with minor allele count (MAC) greater than 20 (genome-wide significance level at p < 5×10^-8^).

In UKB, single variant association analyses identified 89 rare variants (94 variant-phenotype pairs) and 375 common variants (437 variant-phenotype pairs) associated with sleep phenotypes (**Supplementary Table 2 and 3**). Most rare variant associations were linked to the number of unique sleep medications, highlighting a substantial influence of rare coding variation on sleep medication use, potentially reflecting underlying disorder severity and treatment resistance. For sleep disorder diagnoses that were additionally examined in AoU, we identified *MEIS1* associated with restless leg syndrome (**Supplementary Table 4**). In the meta-analyses, we found one variant linked to hypersomnia (*MROH2A*) and sleep apnea (*LMF1*) in European ancestry (Supplementary Table 5) and five single variants in cross-population meta-analyses across all traits (meta-analyzed N = 539,465; **Supplementary Table 6**).

We also conducted rare variant gene-burden analyses for protein-truncating variants (PTVs), damaging missense variants, and synonymous variants using two MAF thresholds (MAF <0.01 and <0.001). We identified 51 genes associated with sleep phenotypes in UK Biobank (p < 2.5×10^-6^; **Supplementary Table 7**), with most of the results observed for PTV burden. Specifically, 6 genes were associated with self-reported phenotypes, *CGGBP1* was associated with post-viral fatigue syndrome, 13 genes were associated with sleep medications with notable *SERPINB6* associated with the number of unique sleep medications, zopiclone as well as any sleep medication, and 26 genes were associated with accelerometer-based measures. In AoU, *CNTLN* was associated with malaise and fatigue and *CXCL6* was associated with post-viral fatigue syndrome in AoU European ancestry, and one gene (*QPCT*) associated with insomnia in AoU admixed American ancestry (p < 2.5×10^-6^; **Supplementary Table 8**).

In total, across both single variant and gene-burden analyses, we identified 260 genes with genome-wide significant associations, representing 309 gene-phenotype pairs implicated in 36 sleep phenotypes (**Figure 2**; **Supplementary Figure 1; Supplementary Table 9**). Specifically, 193 genes were associated with self-reported sleep phenotypes, 12 with sleep disorder diagnoses, 29 with sleep medications, and 28 with accelerometer-based measures. Among these genes, 42 were associated with more than 1 phenotype (**Supplementary Table 10**).

**Figure 2.**
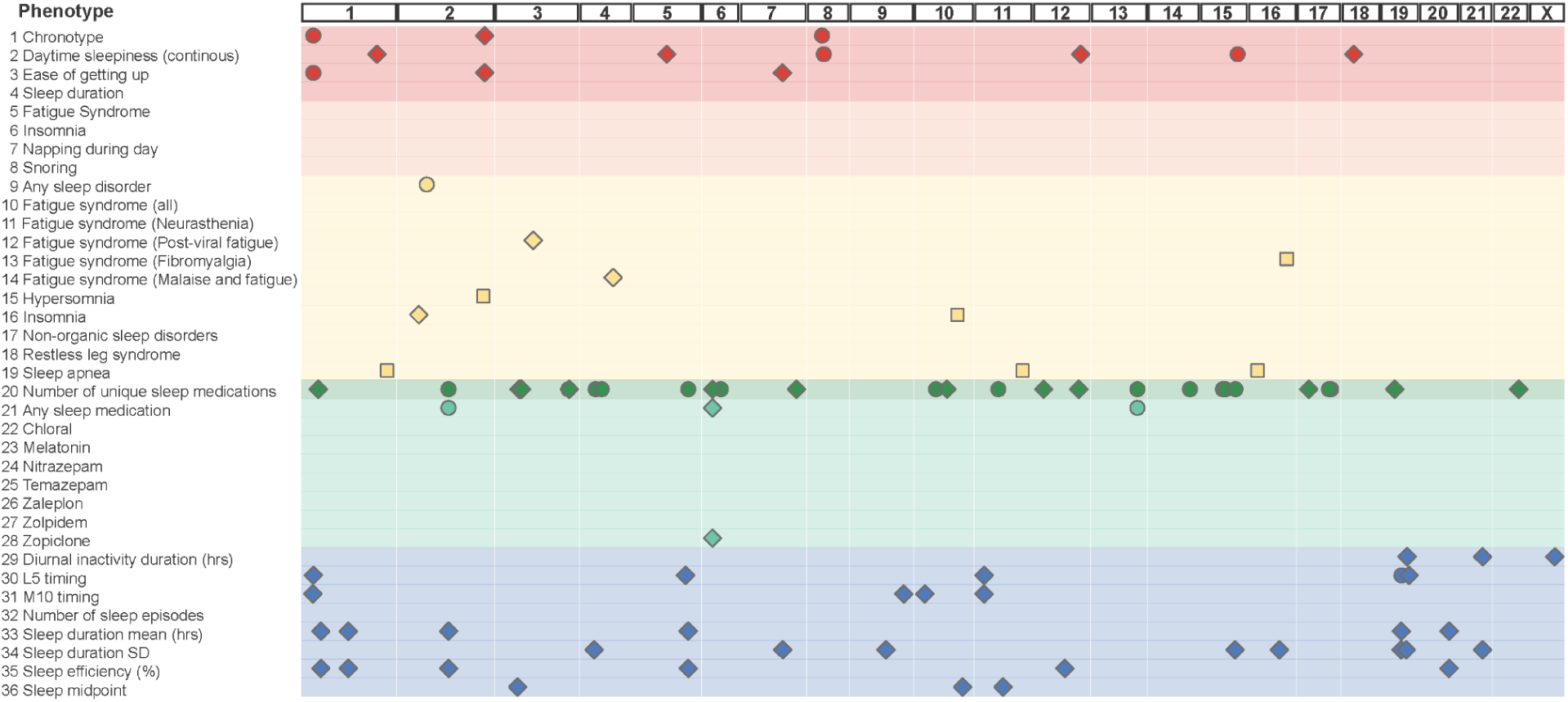
Gene discovery using gene-based, single rare variant and meta-analyzed associations for 36 sleep phenotypes. In total, across both single-variant and gene-based analyses, we identified 260 genes implicated in 36 sleep phenotypes. Specifically, 193 genes were associated with self-reported sleep phenotypes, 12 with sleep disorder diagnoses, 29 with sleep medications and 28 with accelerometer-based measures. This figure includes findings from gene-based analyses (diamonds), rare single variants (circles) and meta-analyses of single variants in the UK Biobank and All of Us (squares).

To identify potential pleiotropic genes across sleep phenotypes, we summarized our gene discovery findings (N _genes_ = 260) across the four major sleep phenotype measurement modalities (**Figure 3a**). Only two genes were found to overlap across measurement modalities. Specifically, we found that *USP34* overlapped between self-reported phenotypes and sleep disorders, while *PER3* was shared between self-reported and accelerometer measures. These findings indicate that exome variant associations are largely specific to the measurement modality with the caveat that the sample size varies across measurement modalities. This suggests that different measurements capture distinct subsets of the sleep phenotype spectrum, as evidenced by greater genetic overlap within measurement modalities than across them.

**Figure 3.**
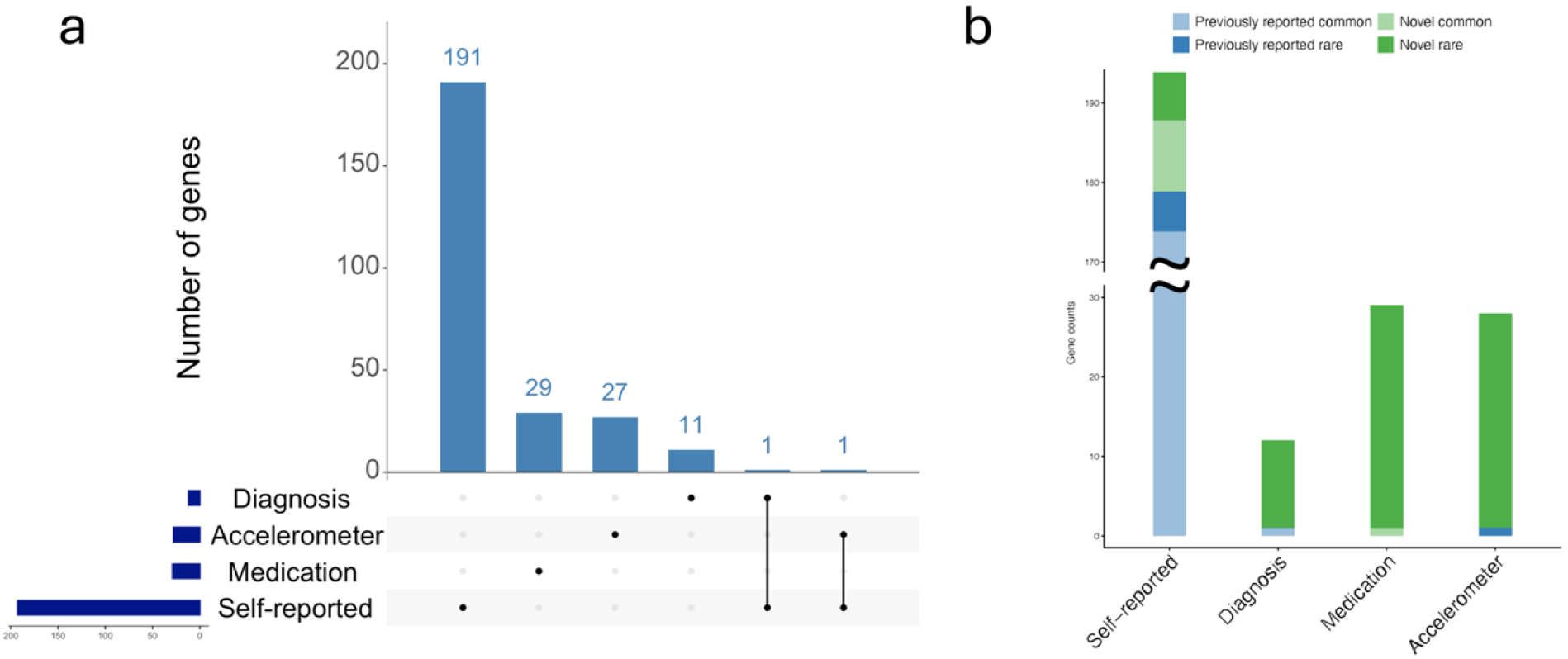
Summary counts of novel loci by measurement modality. (a) Summary of sleep gene findings based on rare and common single variants and gene-based associations across different measurement modalities. (b) Summary of sleep gene findings from the current study compared with previous GWAS and ExWAS studies, with comparisons made within each sleep measure modality. Common and rare groups were defined in the discovery datasets: results from gene-based tests and single variants with MAF < 0.01 were defined as rare, and results from single variants with MAF ≥ 0.01 were considered common.

### Novel Association Signals

To identify previously unreported sleep genes from our discovery findings (N_gene_ = 260), we compared our genome-wide significant associations with all independent genome-wide significant signals reported in 11 recent GWAS ^1,2,10–13^ within each measurement modality, spanning eight self-reported sleep phenotypes, one accelerometer-derived measure, and two sleep disorder diagnoses, as well as with results from a recent ExWAS (**Supplementary Table 11**). Among the 193 genes associated with self-reported sleep phenotypes in our study, 17 genes were not reported previously with self-reported sleep phenotypes, 171 genes overlapped with prior common variant findings, and five genes were reported in previous rare variant exome studies. For sleep disorder diagnoses, the majority of associations were novel, with only *MEIS1* being reported in previous GWAS. As the first whole-exome association study on sleep medications, we reported 28 novel genes from rare variant associations and 1 novel gene (*TTBK1*) from common variant associations. For accelerometer-based sleep measures, 27 additional novel genes were identified from rare variant associations, with only *PER3* being reported in a prior exome analyses (**Figure 3b**).

### MGB Biobank Replication

Replication of significant gene discovery findings was sought using exome sequencing data from 31,275 participants in the MGBB, where 12 matching EHR-based sleep disorders and medication phenotypes were available (**Supplementary Table 1**). After extracting single variants and genes identified in the discovery analysis, we performed annotation using the same approach as in the UKB. Although with limited sample size in MGBB, we still attempted to replicate single variant associations from our discovery analysis for 132 variants with MAC > 5 identified in MGBB. Only 15 low-frequency variants (17 variant–phenotype pairs) with MAF < 0.05 showed nominally significant association (p < 0.05), where the low replication rate for single variant associations is expected due to the limited power in MGBB. Notably, for all single variant associations replicated with nominal significance, consistent direction with the discovery data were shown except for one variant in *PER2* (**Figure 4a**; **Supplementary Table 12**). For gene-burden association replication, of 54 genes identified in discovery analysis, 25 genes showed nominally significant association (p < 0.05) at the MAF threshold of 0.001. For genes associated with EHR-based sleep disorder or medication phenotypes in discovery analysis, where phenotypes were aligned with MGBB phenotypes, all 12 nominal significant tests demonstrated directional consistency in MGBB (**Supplementary Table 13**). Across 125 gene-phenotype tests among all 36 sleep phenotypes tested in discovery analysis, 69 (55.2%) showed concordant effect directions with the discovery dataset (**Figure 4b**).

**Figure 4.**
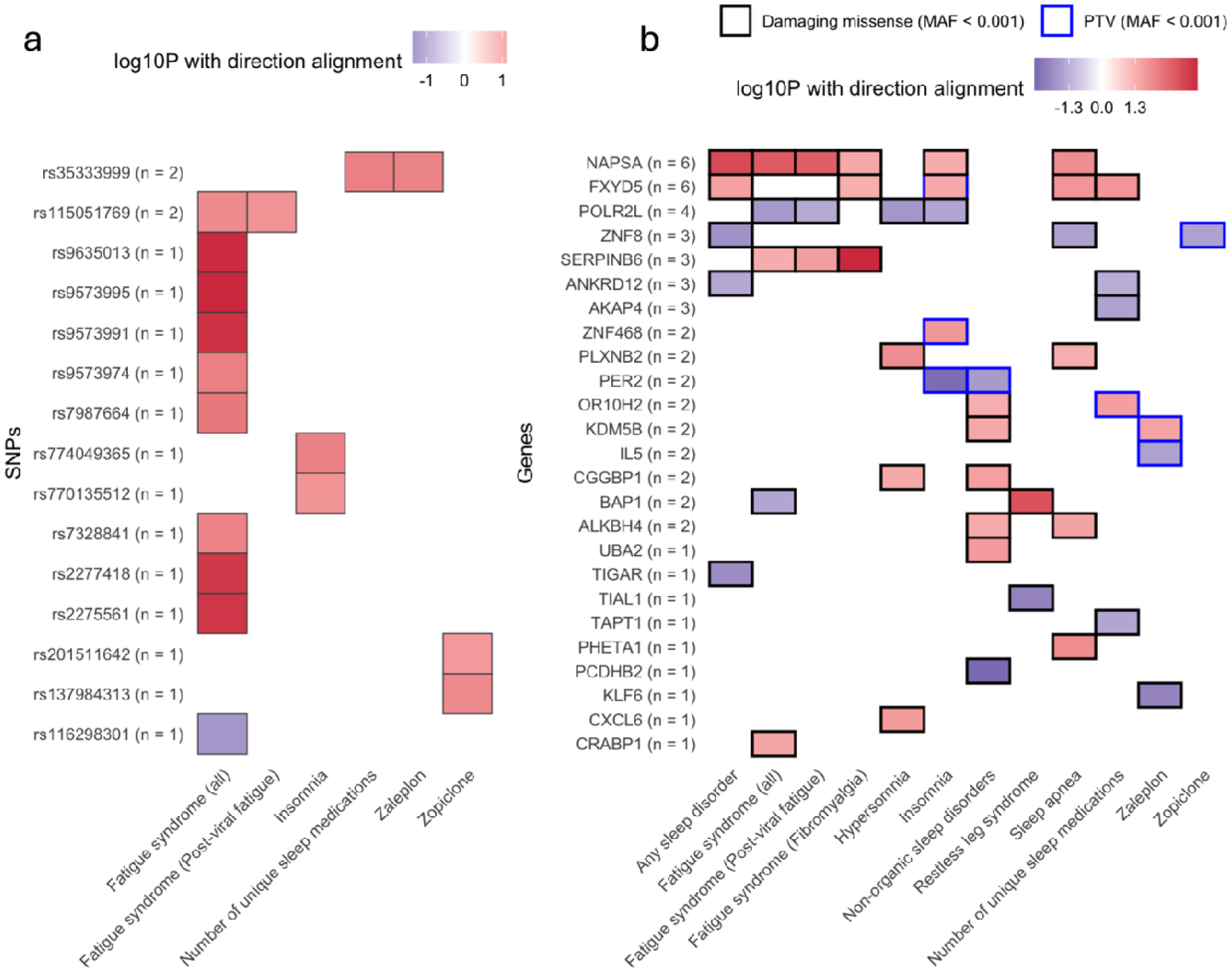
Replication of single-variant associations (a) and ultra-rare variant gene-burden tests (b) in the Mass General Brigham Biobank. Replication of both single-variant associations and ultra-rare (MAF < 0.001) gene-burden tests in the Mass General Brigham Biobank. The x-axis shows phenotypes in the replication dataset; mapping between discovery and replication phenotypes is described in the Methods. The number of nominally significant phenotypes associated with each rare variant (a) or gene (b) is indicated in parentheses.

### Rare Variant Gene-Burden Heritability and Genetic Correlation

Burden Heritability Regression (BHR) quantifies phenotypic variance explained by the burden of rare coding variants.^45^ We applied BHR to estimate burden heritability for 16 sleep-related traits with a genomic control lambda (λ**_GC_**) > 0.7, using recommended stratification by allele frequency and functional categories (Methods).

For aggregated PTVs, all self-reported, diagnosis, and medication sleep traits, as well as two (out of five) accelerometer-based traits exhibited nominally significant (p < 0.05) non-zero burden heritability. Estimates ranged from 0.08% to 0.41% for self-reports, 0.39% for fatigue diagnoses, 0.17% to 0.31% for sleep medication phenotypes, and 0.7% for accelerometer measures (**Figure 5a; Supplementary Table 14**). Damaging missense variants contributed less burden heritability than PTVs across both ultra-rare and rare variant bins. Only self-reported chronotype (h² = 0.19%) and daytime sleepiness (0.08%), along with accelerometer-based diurnal inactivity duration and sleep midpoint (both 0.78%) showed significant (p < 0.05) non-zero heritability for damaging missense variants (**Figure 5b**).

**Figure 5.**
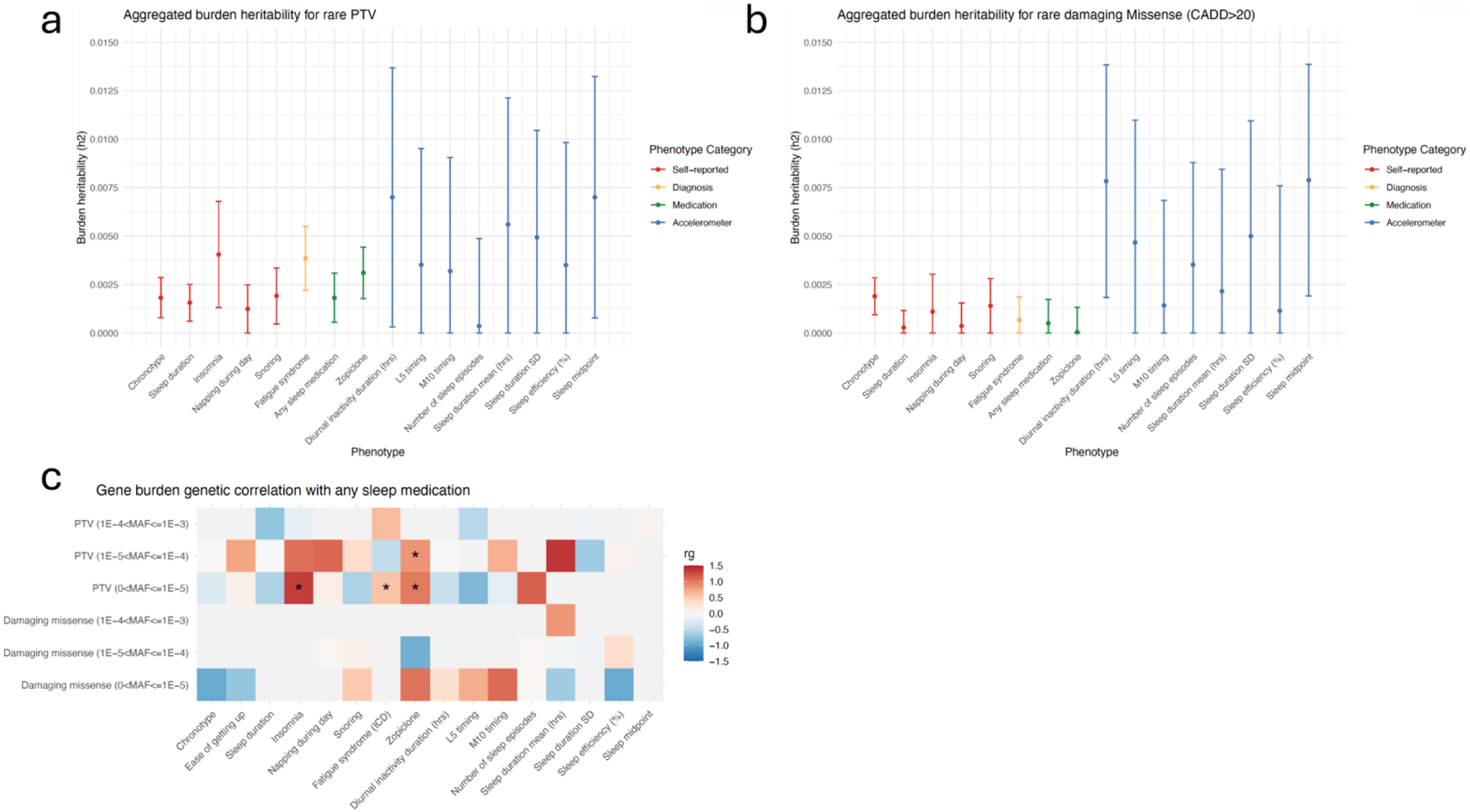
Burden heritability across sleep phenotypes and burden genetic correlation with any sleep medication in the UK Biobank. (a) SNP-based burden heritability for sleep phenotypes from rare protein truncating variants. (b) SNP-based burden heritability for sleep phenotypes from damaging missense variants with CADD≥ 20. (c) Rare gene-burden genetic correlation between any sleep medication and other sleep traits. The sleep phenotypes with lambda less than 0.7 were excluded. CADD: combined annotation dependent depletion.

As the first study to examine the rare variant architecture of sleep medication use, we also used BHR to estimate the burden genetic correlation between the aggregate “any sleep medication” phenotype and other sleep traits (**Figure 5c; Supplementary Table 15**). Any sleep medication showed strong genetic correlation with use of zopiclone for ultra-rare (MAF < 1×10**^-5^**) PTV variants. Specifically, genetic correlation with zopiclone among ultra-rare PTVs was 0.99 and among rare PTVs was 0.84. Notably, self-reported insomnia and fatigue syndrome diagnosis showed significant (p < 0.05) burden correlation with any sleep medication use, indicating a shared rare variant component between sleep problems and and use of non-anxiolytic sleep medications. Although the burden-based genetic correlations for very rare variants (1×10^-4^ > MAF > 1×10^-5^) between medication use and both self-reported and accelerometer-derived sleep phenotypes appeared substantial, they did not reach statistical significance, likely reflecting limited statistical power.

### Network Analyses on Enriched Biological Pathways

To explore the biological mechanisms underlying the sleep phenotype spectrum, we conducted gene-set enrichment and network analyses using ShinyGO^14^ with significant sleep genes. We analyzed genes associated with the number of unique sleep medications and self-reported sleep phenotypes separately to assess whether their underlying pathways were shared or distinct. Using the Gene Ontology (GO) Biological Process gene sets from Ensembl release 92, we mapped 29 genes related to sleep medication (Methods). The most significant and most connected pathway was cerebral cortex tangential migration (FDR-p < 0.05), followed by other fundamental biological processes related to brain development (FDR-p < 0.2) (**Supplementary Figure 2A; Supplementary Table 16 and17**). In contrast, analysis of 193 genes associated with self-reported sleep phenotypes identified 30 pathways (FDR-p < 0.05) predominantly involved in more specific circadian-related processes such as circadian rhythm, rhythmic process, and photoperiodism (**Supplementary Figure 2B; Supplementary Table 18 and 19**). These results suggest that sleep medication-related genes are enriched for general neurodevelopmental pathways, while self-reported sleep traits are linked to more specialized circadian regulation pathways.

### Temporal Expression Trajectories for Sleep Genes

To characterize the trajectory of sleep-related gene expression in the brain across development, we used gene expression data from the BrainSpan Phylogenetic and Ontogenetic dataset^15^ (Methods). This revealed distinct biological signatures for genes related with each sleep phenotype category, highlighting critical windows of gene expression from gestation through maturity (**Figure 6, Supplementary Table 20**). Genes associated with sleep medications demonstrated a sharp, bell-shaped trajectory, with expression increasing steadily throughout the prenatal period to reach a notable peak during the late prenatal stage, followed by a rapid decline into infancy and childhood. In contrast, expression of genes related with self-reported sleep phenotypes maintained high, stable expression throughout the prenatal period and infancy before experiencing a pronounced “dip” during childhood, after which expression levels recovered steadily through adolescence and adulthood. A similar pattern was observed for sleep disorder related genes, which exhibited a consistent downward trend from the early prenatal stage to a minimum in childhood, followed by a marked increase in expression during adulthood. Finally, the accelerometer-derived phenotype genes showed a more linear-style decrease over time; these genes were most active during early brain development and showed a continuous reduction in mean expression as the brain matured, eventually reaching a plateau from childhood to adulthood.

**Figure 6.**
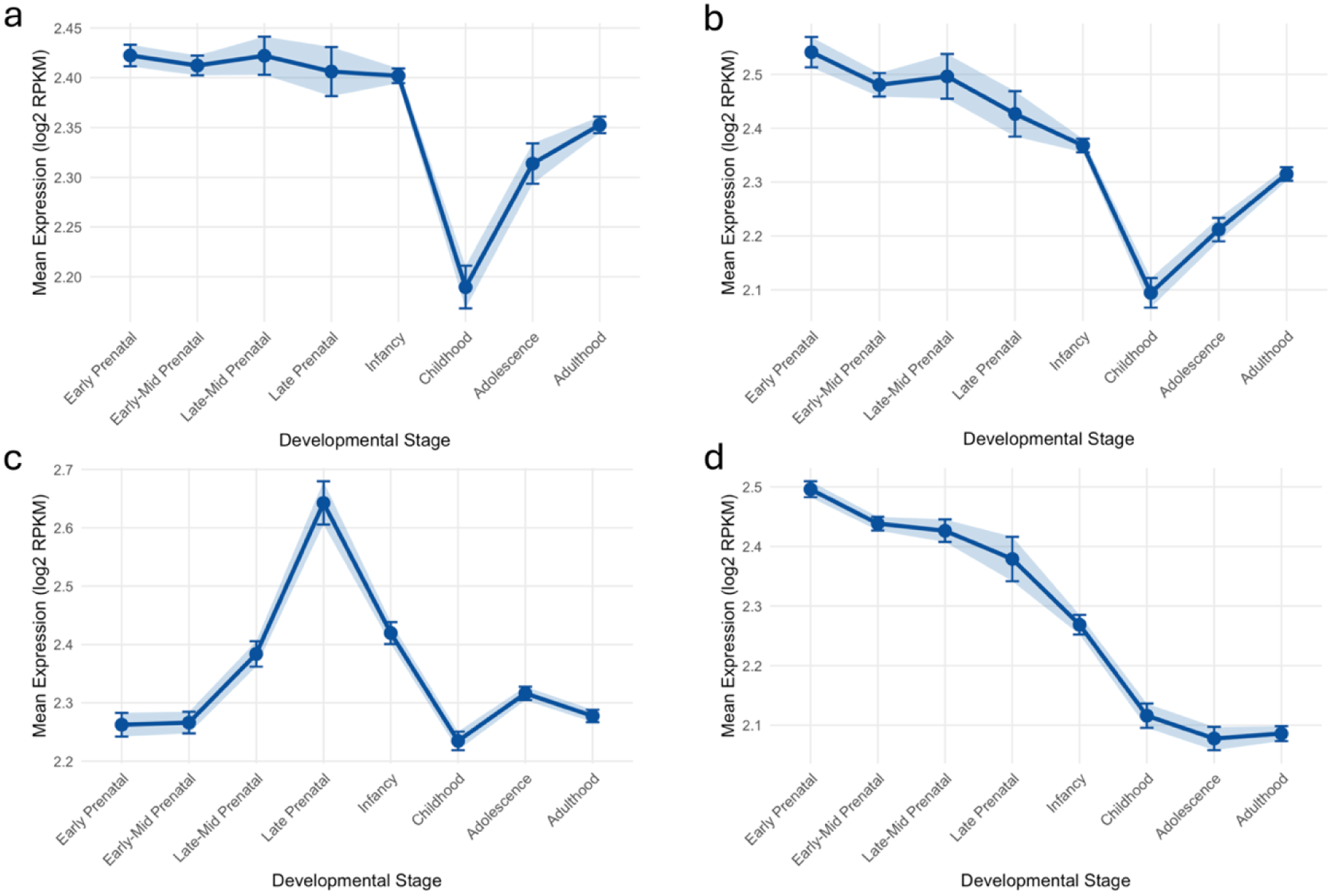
Temporal expression of sleep genes identified from different measurement modalities over the life course in the human brain. Temporal expression trajectories for (a) genes associated with self-reported sleep (N = 184), (b) genes associated with sleep disorders (N = 10), (c) genes associated with sleep medications (N = 27), and (d) genes associated with accelerometer-based sleep measures (N = 25). Samples were split into four prenatal and four postnatal periods, based on whole-brain tissue from the BrainSpan dataset.

## Discussion

This exome-wide association study advances our understanding of the genetic underpinnings of diverse sleep phenotypes, which included a wider spectrum of sleep disorders and a novel focus on sleep medication use based on electronic health records (EHR) from multiple population biobanks (UK Biobank and All of Us Research Program). By analyzing sequencing data across 36 sleep phenotypes, we identified 260 genes associated with sleep phenotypes, including 82 novel genes. Notably, we replicated a specific association of *MEIS1* with sleep disorder diagnoses and discovered 29 genes implicated in sleep medication use. Many of these findings were replicated in an independent biobank (Mass General Brigham Biobank), with largely consistent direction of effect. Burden heritability analyses indicated small contributions from rare protein-truncating and damaging missense variants to most sleep phenotypes. Gene set enrichment analysis showed that genes associated with sleep medications are enriched in biological processes linked to general brain development, whereas genes associated with self-reported sleep phenotypes are more specifically implicated in circadian regulation pathways. Finally, different phenotyping approaches lead to sleep phenotypes exhibiting divergent patterns of gene expression across brain development. These findings highlight the value of expanding the breadth of sleep phenotypes for genetic discovery, with distinct genetic architectures across self-reported, disorder diagnoses, medication use, and accelerometer-based phenotypes, offering insights into biological mechanisms of sleep-related health problems and prioritizing potential therapeutic targets.

This is the first rare variant association study to examine sleep medication usage leveraging the EHR, a phenotype likely capturing both sleep disorder severity and treatment resistance. The number of unique sleep medications is conceptually analogous to treatment resistant depression, schizophrenia or hypertension, which are commonly defined by the number of medications prescribed,^16–18^ yet it is a novel application for sleep medications. Meanwhile, we acknowledge that this sleep phenotype could be often prescribed non-specifically (e.g., for pain, or nonspecific insomnia complaints), and our definition may aggregate heterogeneous underlying pathologies, similar to concerns raised for treatment-resistant depression, potentially diluting effect sizes and reducing power. Nonetheless, this pragmatic, medication-based sleep phenotype offers a scalable starting point for capturing more severe sleep disturbance and motivates future work to refine this phenotype with richer clinical and behavioral data.

In our study, we identified 29 genes related to the number of unique sleep medications, and 24 out of 29 (82.8%) have not previously been implicated in any sleep phenotypes. While five genes (*CD82, FAM114A1, KIAA1143, SPINK8*, and *ZKSCAN3*) overlap with prior GWAS findings, the predominance of novel signals highlights the added value of examining rare coding variation in an expanded phenotypic landscape. Notably, *SERPINB6* showed one of the strongest and most consistent associations across both single-variant and gene-burden analyses. *SERPINB6* encodes an intracellular serine protease inhibitor that regulates immune-related protease activity and modulates neuropsin expression in the brain, ^19,20^ linking immune regulation with synaptic plasticity mechanisms. Moreover, several implicated genes converge on pathways central to neuronal integrity and circuit function. *ASAH1* regulates sphingolipid metabolism critical for neuronal survival and stress responses,^21^ *KDM5B* modulates neurodevelopmental gene expression through histone demethylation,^22,23^ and *RASGRF1* contributes to Ras-dependent synaptic and circadian signaling.^24^ Collectively, these findings suggest that rare coding variants affecting neurodevelopment, synaptic plasticity, and network stability may contribute to sleep disorder burden and treatment complexity, extending the genetic architecture of sleep phenotypes beyond common variant GWAS and highlighting potential targets for drug development.

Beyond novel gene discovery, ExWAS brings in new insights to our understanding of the shared genetic underpinning of sleep phenotypes, especially compared with previous GWAS. For example, *PER2* is associated with chronotype and daytime sleepiness in UKB (PTV burden leads to morning chronotype and easier to get up in the morning) which is consistent with a previous ExWAS report that also leveraged UKB. However, *PER2* PTV burden was associated with higher risk for insomnia and non-organic sleep disorders in MGB, albeit with only a nominally significance. Intriguingly, while previous GWAS showed a negative genetic correlation between insomnia and morning chronotype as well as insomnia and ease of getting up,^3^ previous ExWAS results indicated a positive loss of function rare variant burden genetic correlation between insomnia risk and morning chronotype as well as ease of getting up.^6^ Notably, these rare variant burden genetic correlation findings are concordant with our *PER2* associations observed in UKB and MGBB. Given the well-established role of *PER2* in sleep regulation,^3,13,25^ the contrasting genetic correlation patterns suggest that rare and common variants may exert distinct effects on these traits. Together, these findings indicate that both genome-wide variation and rare variation within *PER2* itself may differentially contribute to chronotype-related phenotypes and insomnia risk, which could be a future direction when larger samples become available.

Most self-reported sleep phenotypes, fatigue syndrome, and sleep medication use, showed significant burden heritability driven by rare protein-truncating variants (PTVs), with a weaker contribution from damaging missense variants. In contrast, accelerometer-based measures like diurnal inactivity duration and sleep midpoint exhibit higher heritability from damaging missense variants than from PTVs. This suggests potential distinct biological mechanisms underlie different sleep phenotype measurement modalities that capture different aspects (e.g., more specific parameters reflected in the accelerometer-based measures or broader self-reported phenotypes).^1,26^ Furthermore, we observed strong rare gene-burden genetic correlations between the use of any sleep medication and zopiclone as well as with self-reported insomnia and fatigue diagnoses. This pattern highlights a close genetic relationship within sleep medication use traits and aligns with clinical observations that sleep medication is predominantly prescribed for insomnia symptoms.^27,28^ The link between sleep medication use and fatigue syndromes may be due to shared biological pathways acting through neurotransmitter systems of GABA and serotonin,^29,30^ or the moderately strong genetic correlation between insomnia and fatigue.^11,13^

We observed enrichment of broad biological process pathways (e.g., cell development, nervous system development, cell differentiation, neurogenesis) for genes associated with sleep medication use. This indicates potential shared biological pathways for sleep medication use and neuropsychiatric problems.^31^ In contrast, genes related to self-reported sleep phenotypes showed pathway enrichment more specifically tied to circadian rhythm. Notably, prior GWAS of self-reported traits highlighted broader pathways,^11,32^ suggesting that ExWAS may better resolve directly relevant biological mechanisms. In addition, the distinct brain trajectories also indicated distinct developmental expression patterns for different sleep measurement sources. Genes associated with sleep medication mostly for insomnia treatment peaked during the late prenatal phase, aligning with the critical formation of neurotransmitter systems that serve as primary targets for sleep-modulating drugs.^33^ Conversely, the self-reported and sleep disorders associated genes exhibit a “U-shaped” expression trajectory, with a low point in childhood. This dip occurs during a window of intense synaptic pruning and homeostatic sleep refinement, suggesting these genes regulate the transition from early developmental plasticity to adult sleep-wake stability.^34^

This study has several limitations. First, analyses were largely restricted to individuals of European ancestry due to limited availability of exome data from other populations. We partially mitigate this limitation by including African American and Latino American samples from AoU; however, larger sequencing studies in non-European samples are warranted to further expand our understanding in rare variant genetic architecture for sleep phenotypes. Second, potential overlap between MGBB and AoU may have inflated replication signals; however, most of the replicated findings were driven by discoveries in the UKB. Third, although the number of sleep medications as a sleep phenotype showed high statistical power for genetic discovery, it was broadly defined, which may obscure biology specific to individual medications.

This study also offers several notable strengths. First, it is the first ExWAS to systematically leverage EHR-derived and accelerometer-based sleep phenotypes alongside self-reported phenotypes, enabling both clinical insight and cross-measurement comparisons. Second, we conducted meta-analyses across multiple biobanks and diverse populations to enhance generalizability and statistical power. Third, replication of key findings in an independent biobank supports the robustness of our results. Overall, this study provides novel insights into the role of rare genetic variants in the etiology of sleep phenotypes, particularly in relation to sleep medication use at the population level.

## Online Methods

### Ethics

The UK Biobank (UKB) received ethical approval from the North West Multi-centre Research Ethics Committee (https://www.ukbiobank.ac.uk/learn-more-about-uk-biobank/about-us/ethics). The current study was conducted under UKB application number 26041. The data in the UKB were collected with written informed consent obtained from all participants. The All of Us (AoU) Research Program protocol was approved by the Institutional Review Board (IRB) of the All of Us Research Program. The All of Us IRB follows the regulations and guidance of the NIH Office for Human Research Protections for all studies, ensuring that the rights and welfare of research participants are overseen and protected uniformly. Adults 18 years and older who have the capacity to consent and reside in the USA or a US territory at present are eligible. Informed consent for all participants is conducted in person or through an eConsent platform that includes primary consent, HIPAA Authorization for Research use of EHR and other external health data, and Consent for Return of Genomic Results. The Human Research Committee of Mass General Brigham (MGB) approved the MGB Biobank (MGBB) research protocol (no. 2009P002312). The data in the MGBB were collected with written informed consent obtained from all participants.

### Sleep phenotypes in UKB

We assessed sleep phenotypes across four categories: self-reported measures, sleep disorder diagnoses based on International Classification of Diseases (ICD) codes from electronic health records (EHR), medication prescription records from EHR, and accelerometer-based measures, which provide more detailed information but are only available for a smaller number of participants. **Supplementary Table 1** provides further information on the coding and sample size of each phenotype. Continuous phenotypes were rank-based inverse normal transformed to improve normality.

Self-reported sleep questionnaires were administered to UKB participants at baseline assessment. We used the following 8 self-reported sleep phenotypes in our analysis: 1) chronotype was defined as a continuous variable generated by a morning versus evening person, where 1 is “definitely a morning person” and 4 is “definitely an evening person”. 2) daytime sleepiness was derived based on the question “How likely are you to doze off or fall asleep during the daytime when you don’t mean to? (e.g. when working, reading or driving)?” as a continuous variable. 3) ease of getting up in the morning was treated as a continuous variable ranging from 1 to 5, derived from the question “On an average day, how easy do you find getting up in the morning?” with 1 representing very easy. 4) sleep duration was treated as a continuous variable. Participants were asked “About how many hours sleep do you get in every 24 h? (please include naps)”, with responses in hour increments. Extreme responses of less than 3 h or more than 14 h were excluded. 5) self-reported fatigue syndrome was treated as a binary variable based on the question “Have you ever been told by a doctor that you have had chronic fatigue syndrome or myalgic encephalomyelitis?”. 6) self-reported insomnia was treated as a binary variable, comparing those who answered “Never” versus those who answered “Usually”, and excluding those who answered “Sometimes”. 7) daytime napping was treated as a binary variable based on the question “Do you have a nap during the day?”. An answer of “Usually” was coded as “Yes” and “Sometimes” or “Never/rarely” was coded as “No”. 8) snoring was also treated as a binary variable with yes versus no answers for the question “Does your partner or a close relative or friend complain about your snoring?”. For all phenotypes, answers of “Do not know” or “Prefer not to answer” were coded as missing.

For EHR-based sleep phenotyping, we extracted data from both primary care (GP; General Practitioner) and secondary care in-patient (HES; Hospital Episode Statistics) data. We mapped READ2, CTV-3, BNF, DM+D and ICD 9 codes to ICD 10. For each of the following sleep disorders, cases were defined as those with at least one ICD10 code in their EHR. Fatigue syndrome was identified from several different ICD code sources, including neurasthenia (F48.0), postviral and related fatigue syndromes (G93.3), fibromyalgia (M79.7), and malaise and fatigue (R53). In addition to the composite “fatigue syndrome” phenotype, each of the 4 disorders were also analyzed as distinct phenotypes. Hypersomnia cases were identified from F51.1 and G47.1. Insomnia cases were defined as any diagnosis of F51.0 or G47.0. Non-organic sleep disorders were identified through sleepwalking (F51.3), sleep terrors (F51.4), nightmare disorders (F51.5) and parasomnia that involves abnormal behaviors or experiences (G47.5). Restless leg syndrome cases were identified by specified extrapyramidal and movement disorders (G25.8). Sleep apnea was defined using G47.3, which includes obstructive, central, and mixed forms of sleep apnea. A composite “any sleep disorder” phenotype was also defined, with cases having at least 1 recorded ICD code from any of the above disorders.

Based on both primary and secondary care data records, we extracted medication records on sleep disorder-related prescriptions, including circadin, chloral, clomethiazole, flurazepam, melatonin, nitrazepam, nytol, promethazine, rohypnol, temazepam, triazolam, zaleplon, zileze, zolpidem, and zopiclone, while excluding medications that are also commonly used for anxiety disorders. Two composite phenotypes were derived based on these drugs: 1) “number of unique sleep medications” is a continuous variable which counts the number of unique sleep medications a patient has been prescribed. 2) “any sleep medication” is a binary indicator for a prescription history for any sleep-related medication. Each medication with more than 100 prescribed patients was also treated as a distinct phenotype, including chloral, melatonin, nitrazepam, temazepam, zaleplon, zolpidem, and zopiclone.

Accelerometer data was available for 103,711 UKB participants who participated in a follow-up study between 2.8 and 9.7 years after study baseline. A triaxial accelerometer device (Axivity AX3) was worn by participants for a continuous period of up to 7 days. We used the same quality control and processing pipeline to derive accelerometer-based sleep traits as described in previously published GWAS,^1^ excluding days with <16 hours of valid wear time. We first defined the following sleep characteristics, which we then used to create 8 sleep phenotypes. Sleep period time window (SPT-window) was defined by using a validated algorithm based on changes in wrist z-angle across 5-min rolling windows. Periods of inactivity lasting ≥30 min were identified, and those <60 min apart were combined into blocks, with the longest block defining SPT-window. Sleep episodes within the SPT-window were defined as ≥5 min of minimal movement (z-angle change ≤5°). We then created 8 phenotypes related to sleep timing, duration and quality: diurnal inactivity, least-active 5 hours (L5) timing, most-active 10 hours (M10) timings, number of nocturnal sleep episodes, total sleep duration in hours, sleep duration variability, sleep efficiency, and sleep midpoint. Diurnal inactivity was the total duration of inactivity bouts occurring outside the SPT-window, capturing restful but not sedentary behavior like sitting or watching TV. For timing related traits, least-active 5 h (L5) and most-active 10 h (M10) timings were defined as the midpoints of the respective periods with lowest and highest average activity, based on rolling 5-h and 10-hour windows. The number of nocturnal sleep episodes was the count of episodes within the SPT-window; individuals with ≤5 or ≥30 average episodes were excluded. Total sleep duration was calculated by summing sleep episodes within the SPT-window. Individuals averaging <3 hours or >12 hours of sleep were excluded. Sleep duration variability (standard deviation) was analyzed for individuals with 7 days of data. Sleep efficiency was calculated as sleep duration divided by the SPT-window duration. Sleep midpoint was calculated as the midpoint between the first and last sleep episodes within the SPT-window, expressed as hours since the previous midnight.

### The UKB whole-exome sequencing data

Whole-exome sequencing (WES) data for UK Biobank participants was generated by the Regeneron Genetics Center on behalf of the UKB Exome Sequencing Consortium.^35^ Sequencing was performed using Illumina NovaSeq 6000 platforms with xGen Exome capture kits. Reads were aligned to the GRCh38 reference genome using Burrows-Wheeler Aligner-MEM v0.7.17.^35,36^ Single-nucleotide variants and indels were identified by generating gVCF files with WeCall v1.1.2, followed by joint genotyping with GLnexus v0.4.0.^37^ The resulting VCF underwent quality control through the Regeneron Genetics Center’s ‘Goldilocks’ pipeline. As of November 2020, 454,787 participants had QC-passed WES data available for approved researchers on the UKB Research Analysis Platform.

Variants were annotated with Variant Effect Predictor (VEP) v96^38^ and mapped to GENCODE release 30 canonical transcripts.^39^ Protein truncating variants (PTVs), including stop-gain, splice-disruptive, and frameshift events, were assessed using LOFTEE (a VEP plugin),^40^ and high-confidence PTV variants were retained. Missense variants with CADD scores above 20 were considered deleterious.

Imputed genotype data, filtered for quality, was obtained for all UKB participants. Genotypes were imputed using Haplotype Reference Consortium,^41^ UK10K,^42^ and 1000 Genomes Project reference panels,^43^ resulting in over 90 million variants. Quality control excluded variants with imputation INFO scores below 0.8 and minor allele frequencies (MAF) below 0.01 using PLINK v2.^44^ Additionally, individuals with inconsistent reported and genetic sex, sex chromosome aneuploidies, or UKB withdrawals (as of August 24, 2020) were removed from the dataset.

### Sleep disorder phenotypes in All of Us

Sleep phenotypes were identified from phecodes in the linked EHR, mapped to the same ICD code inclusion criteria used in UK Biobank (**Supplementary Table 1**). To leverage the diverse ancestry samples in All of Us, analyses included the three population groups, including European (EUR), African American (AFR), and Latino American (AMR). For individuals of European ancestry, 8 phenotypes were analyzed: fatigue syndrome (from three sources: post-viral and related fatigue syndromes, fibromyalgia and malaise and fatigue), hypersomnia, insomnia, non-organic sleep disorders, restless leg syndrome, and sleep apnea. For African American and Latino American groups, 6 phenotypes with sufficient case numbers were included in cross-population meta-analyses: fatigue syndrome (from two sources: fibromyalgia and malaise and fatigue), hypersomnia, insomnia, restless leg syndrome, and sleep apnea.

### Whole-exome data processing in All of Us

We performed quality control (QC) on genotype data from both the Allele Count/Allele Frequency (ACAF) threshold callset and the exome callset^45^ in the AoU Workbench. Variants were restricted to those that passed Variant Quality Score Recalibration (VQSR) criteria (based on the ‘FT’ field) and had global allele counts (AC) > 0. Genotypes were further filtered to retain only those with genotype quality (GQ) ≥ 30 and minor allele balance > 0.2 for all alternate alleles in heterozygous genotypes. At the sample level, we first applied hard filters to exclude individuals who were older than 100, had undefined sex, lacked PCA-defined ancestry in the AoU table, or were among 570 duplicates identified from kinship scores in the AoU relatedness table.^46^ To further minimize stratification, we reran PCA within each ancestry group on the filtered set and removed outliers based on multidimensional distances from ancestry-specific PC centroids (described below). After these steps, ∼90% of samples (214,216) passed QC.

Genetic ancestry is defined in the same way as the ancestry groups assigned by PCA from the original AoU data. To reduce stratification and enable robust association analyses, we performed a process of removing outliers with a larger centroid distance computed from ancestry-specific PC scores within each pre-specified ancestry group. Here, to further refine ancestry classifications, our goal was to prune ancestry outliers within assigned labels based on subcontinental structure. Using those LD-pruned variants, we then reran PCA among samples within each assigned genetic ancestry label using individuals determined to be unrelated in the AoU relatedness information table.

All exome single variants were annotated using the Ensembl VEP, which predicts the molecular consequences of each variant on overlapping gene transcripts based on the GRCh38 reference genome. Variants were categorized into four categories. First, predicted Loss of Function (pLoF): Variants likely to disrupt gene function, including stop-gain, frameshift indels, and splice-site disrupting variants, considered equivalent to PTVs in the UK Biobank dataset. Second, missenseLC (Low Confidence pLoF): Variants causing amino acid substitutions predicted to be functionally damaging but with lower confidence on loss-of-function impact. These are identified as damaging missense variants in the UK Biobank analyses. Third, synonymous variants that do not alter amino acid sequences. Fourth, other variants that fall into all other categories.

### Single variants and gene-set based rare coding variant burden test

We performed single variant association analyses using whole-exome sequencing data from the UKB and AoU to investigate rare coding variant associations with 36 sleep-related phenotypes in UKB and up to 8 sleep disorder diagnoses in AoU based on the sample size across ancestry groups. To maximize statistical power, we conducted meta-analyses of single variant results for up to 8 sleep disorder diagnoses across both cohorts. Gene-based burden tests for PTV and damaging missense variants were carried out independently in UKB and AoU. In UKB, association tests were performed using REGENIE v3.1, with step 1 fitting a ridge regression model (using genotyped variants with MAF ≥1%, call rate >90%, and Hardy–Weinberg equilibrium p > 10^-15^) and leave-one-chromosome-out (LOCO) cross-validation to generate polygenic predictions. Step 2 conditioned single exome variant association tests and gene-based burden analyses on LOCO predictions and covariates. Gene-based burden tests aggregated rare PTVs or damaging missense variants per gene (MAF < 0.01 or MAF < 0.001). All analyses were adjusted for sex, age, age^2^, sex-by-age interaction, sex-by-age^2^ interaction, top 20 PCs, and recruitment center.

In AoU, whole-exome single variant and gene-based burden association analyses for up to 8 sleep disorder phenotypes were performed using a two-step generalized linear mixed model pipeline within SAIGE. Inverse-variance weighted meta-analyses of single variant results were conducted for 8 sleep disorder diagnoses in UKB and AoU European ancestry populations, and for 6 sleep disorder diagnoses across UKB European and AoU European, African, and Admixed American ancestry groups to enhance generalizability. All analyses were adjusted for sex, age, age^2^, sex-by-age and sex-by-age^2^ interaction, and top 20 PCs.

To compare our discovery findings for both single-variant and gene-based results with existing literature on the genetic basis of sleep phenotypes, we grouped the associated genes according to different sleep measurement sources: self-reports, diagnoses, medications and accelerometer-based measures. For each category, we classified associated genes as novel or previously reported. Novel genes are those not reported before within the same sleep measurement source, while previously reported genes have been documented in prior genetic studies for that specific category. Definitions of common and rare variants were based on minor allele frequencies with a threshold at 0.01 within our discovery datasets. Gene burden-based findings were considered rare variant associations. This categorization allows a systematic evaluation of our findings in the context of previous genetic knowledge across diverse sleep phenotypes.

### Replication analyses in Mass General Brigham Biobank

Replication of genetic association findings in UKB and AoU was conducted using an independent whole-exome sequencing study from MGBB, a hospital-based biobank collecting blood samples, electronic medical records, lifestyle, and family history data from approximately 80,000 consented participants (as of November 2021).^47^ We extracted a total of 15 sleep disorder diagnoses and medication records from the Mass General Brigham Biobank (MGBB) based on their EHR data, aligning with the phenotypes used in the discovery datasets. Binary variables were created for any sleep disorder involving the following diagnoses: fatigue syndrome (including postviral and related fatigue syndromes with ICD-10 code G93.3 and fibromyalgia with ICD-10 code M79.7), each individual fatigue syndrome diagnosis, hypersomnia, insomnia, non-organic sleep disorders, restless leg syndrome, and sleep apnea. For medications, the number of unique sleep medications prescribed to each patient was treated as a continuous variable. Additionally, individual medications with more than 200 cases, including zaleplon and zopiclone, were analyzed as separate binary phenotypes.

To identify the MGBB samples of European ancestry for replication, we performed principal component analysis (PCA) based population assignment and PCA in the analytical samples. To this end, we first performed quality control (QC) on the genotype from MGBB, which included 24,787 samples genotyped in two batches: the first with the Multi-Ethnic Genotyping Array (MEGA; 1,416,020 variants) and the second with the Expanded MEGA (MEGA Ex; 1,741,376 variants). The QC procedure followed the MGBB genotype QC pipeline (https://github.com/Annefeng/PBK-QC-pipeline) using PLINK1.9, R, and Python. Variants with call rates >95% and samples with call rates >98% were retained. Variants with batch-specific missing rate differences >0.75% were filtered out. After merging batches, duplicated variants, monomorphic variants, and those not confidently mapped to chromosomes were removed. For population assignment, the genotype data were first combined with 1000 Genomes Project (1KG) phase 3 data, retaining overlapping, bi-allelic, strand-unambiguous variants with MAF ≥5% and call rate ≥98%. Then PCA was performed on the combined study and 1KG reference samples with LD-pruned independent variants (R² < 0.1, window size 200 kb) excluding known long-range LD regions (chr6: 25-35 Mb and chr8: 7-13 Mb). Population assignment employed a random forest classifier trained on the top 6 PCs of 1KG samples, classifying MGBB samples into five populations, i.e., European (EUR), East Asian (EAS), African (AFR), American (AMR), and South Asian (SAS), with prediction probability >0.8. The European subset included 17,287 (69.7%) samples. Within Europeans, samples with mismatched sex, extreme heterozygosity (>5 SD from mean), or relatedness (pi-hat > 0.2) were removed. Variants showing significant batch effects (p < 1.0×10-4) were also excluded. PCA was redone on the remaining 16,701 EUR samples, removing 73 outliers (>6 SD from mean in top 10 PCs). The final in-sample PCs were used to adjust for population stratification in replication analyses.

Whole-exome sequencing (WES) in MGBB was performed with a custom TWIST Biosciences exome panel and sequenced on Illumina NovaSeq with 150 bp paired-end reads, achieving ≥20X coverage for >85% of exonic targets. Variant calling used GATK’s HaplotypeCaller in gVCF mode jointly applied across 26,421 samples. QC on WES data was conducted using Hail v0.2. Autosomal VCFs were imported and merged into a matrix table. Variant-level filtering retained bi-allelic variants with genotype quality (GQ) > 20, call rate > 90%, allele count > 0, mean depth (DP) between 10 and 200, and allele balance (AB) > 0.9. SNPs passed further hard filters including QualByDepth (QD) ≥ 2, FisherStrand (FS) ≤ 60, StrandOddsRatio (SOR) ≤ 3, RMSMappingQuality (MQ) ≥ 40, MQRankSum ≥ −12.5, and ReadPosRankSum ≥ −8. Indels passed the criteria of QD ≥ 2, ReadPosRankSum ≥ −20, FS ≤ 200, and SOR ≤ 10. For the replication, variants and genes identified in discovery datasets were extracted and annotated following the same methods as UK Biobank. Association testing used Firth’s logistic regression for single variants and gene burdens (PTV and damaging missense gene-set burden test on both MAF < 0.01 and MAF < 0.001 thresholds).

We regressed case-control status on variant carrier status, adjusting for the top 20 PCs, mean-centered age, sex, age², sex-by-age interactions, and sex-by-age^2^ interaction. To assess directional consistency between nominally significant (p < 0.05) results in MGBB and significant findings from the discovery dataset, we first reverse-coded effect estimates for sleep duration across single variants and gene-set burden tests, aligning shorter sleep duration with higher risk of sleep disorders. We then compared the direction of effect estimates for each single variant (with aligned effect alleles) and gene-set between MGBB and the discovery data.

### Burden heritability and burden genetic correlation

We used BHR^48^ (https://github.com/ajaynadig/bhr) to estimate the heritability (h**^2^_bhr_**) explained by the gene-wise burden of rare coding variants across sleep traits. As recommended, the variants were stratified into bins according to allele frequency and functional categories. Ultra-rare was defined as MAF < 1 × 10^−5^, and rare was defined as 1 × 10^-3^ > MAF ≥ 10^-5^. Functional annotations of the variants were described in the sections above. All BHR analyses were run with the baseline file based on gnomAD.^38^ The variant-level summary statistics derived from REGENIE outputs were used. Univariate BHR analysis was performed to estimate the h**^2^_bhr_**of each sleep trait, and we aggregated ultra-rare and rare burden heritability for both PTV and damaging missense variants. To better understand the relationship between sleep medication phenotypes and other available sleep related phenotypes, bivariate BHR analysis was then used to compute the burden genetic correlation (r_bhr_). The burden genetic correlations for ultra-rare and rare PTV and damaging missense variants between any sleep medication and other sleep traits were estimated.

### GO Biological Process gene-set enrichment and network analyses

We conducted gene-set enrichment and network analyses using the ShinyGO web tool (version 0.82 based on Ensembl release 92)^14^ to explore the GO biological process pathways enriched among gene sets related to sleep traits, specifically the number of unique sleep medications and aggregated self-reported sleep phenotypes. We included the genes indicated from each measurement modality in the gene list for enrichment and network analysis.

To begin, we input our lists of genes separately into the ShinyGO online interface by pasting gene IDs, which were automatically mapped to Ensembl gene identifiers. ShinyGO uses annotation data primarily from Ensembl and STRING-db to perform enrichment analyses, with the default background comprising all protein-coding genes. For gene-set enrichment, we selected the Gene Ontology (GO) Biological Process category including 15,796 gene sets to identify significantly enriched pathways in our gene list. ShinyGO employs the hypergeometric test to compute p-values for enrichment, with multiple testing correction applied using the Benjamini-Hochberg false discovery rate (FDR) method. Fold enrichment scores are calculated as the ratio of the percentage of genes in the user list within a pathway to the percentage of genes in that pathway from the background set. We then constructed an enrichment network where individual nodes represent enriched functional categories, while edges connect nodes that share a significant portion of overlapping genes. The node size reflects the number of genes within each category, and the node color intensity represents the level of FDR-corrected statistical significance. Edge thickness indicates the degree of overlap between connected pathways, allowing for the identification of clusters of related biological processes.

### Temporal gene expression Trajectory Analysis

To characterize the temporal expression trajectories of the significant genes identified for each measurement modality (N_self-reported gene_ = 184; N_sleep disorder gene_ = 10; N_sleep medication gene_ = 25; N_accelerometer gene_ = 27), we mapped aggregated gene expression data across the human lifespan using the BrainSpan Phylogenetic and Ontogenetic dataset.^34^ Whole-brain expression levels were averaged at each of the eight discrete developmental phases: Early Prenatal, Early-Mid Prenatal, Late-Mid Prenatal, Late Prenatal, Infancy, Childhood, Adolescence, and Adulthood. By grouping the data by these specific time points, we were able to visualize longitudinal trajectories and identify the stages of brain development where these gene sets are most biologically active.

## Code availability

All code developed to process genetic data and subsequently develop PRS models are available via Zenodo. REGENIE (v3.1), SAIGE (v1.3.0) and R (v4.5) were the predominant tools used in this study.

## Data Availability

The exome single-variants and gene-based results from the UK Biobank for each phenotype and the meta-analyses for single-variants are available on GWAS Catalog.

## Acknowledgements

This research has been conducted using the UK Biobank Resource under Application Number 26041. We thank the participants of UK Biobank. We gratefully acknowledge All of Us participants for their contributions, without whom this research would not have been possible. We also thank the National Institutes of Health’s All of Us Research Program for making available the participant data examined in this study. We acknowledge the Mass General Brigham Biobank for providing samples, genomic data, and health information data. W.L., and K.J.K. are supported by 1R01HL179112. T.G. is supported by R01HG012354 and R01MH130899. H.M.O is supported by R01HG012810.

## Author contribution

R.S., H.M.O., J.L., and C.-Y.C., conceived and supervised the study. C.-Y.C., Y.Z and W.L. performed the analyses. K.K. supervised the analyses in the All of Us Research Program. L.K., S.J., M.M., and J.V. performed phenotype data QC analysis. C.-Y.C. and Y.Z wrote the manuscript. A.W., M.W., J.T., T.G., and H.T. critically revised the manuscript. All authors reviewed and approved the final version of the manuscript.

## List of supplementary tables

**Supplementary Table S1.** Description of sleep phenotypes included in the study, including LambdaGC for exome-wide single variant association analysis

**Supplementary Table S2.** Significant single rare variant (MAF<0.01, MAC>=20) association (P<5×10-8) in samples of European ancestry in UKB

**Supplementary Table S3.** Significant single common variant (MAF>=0.01) association (P<5×10-8) in samples of European ancestry in UKB

**Supplementary Table S4.** Significant single common variant association (P<5×10-8) in All of Us European ancestry samples

**Supplementary Table S5.** Significant (P<5×10-8) single variant association from meta-analysis of phecodes in samples of European ancestry

**Supplementary Table S6.** Significant (P<5×10-8) single variant association from cross-population meta-analysis of phecodes

**Supplementary Table S7.** Significant exome-wide gene-based PTV and damaging missense burden association (P<2.5×10-6) in samples of European ancestry in UKB

**Supplementary Table S8.** Significant exome-wide gene-based PTV and damaging missense burden association (P<2.5×10-6) in samples of European ancestry in All of Us

**Supplementary Table S9.** Summary of genes associated sleep phenotype with cross-phenotype Bonferroni correction (N = 260)

**Supplementary Table S10.** Summary of sleep phenotype associated genes (corresponding to Supplementary Table 9)

**Supplementary Table S11.** Summary of previous sleep related GWAS

**Supplementary Table S12.** Single variants significant findings replication in MGBB

**Supplementary Table S13.** Gene-based significant findings replication in MGBB for each distinct gene-phenotype pair

**Supplementary Table S14.** Rare variant burden heritability estimate based on Burden Heritability Regression (retained only phenotypes with rare single variant association lambdaGC > 0.8)

**Supplementary Table S15.** BHR genetic correlation with any medication, based on rare PTV and damaging missense

**Supplementary Table S16.** Enrichment of self-reported sleep genes in Go Biological Process pathways

**Supplementary Table S17.** Enrichment of the number of unique sleep medications genes in Go Biological Process pathways

**Supplementary Table S18.** Edges (gene overlaps) between enrichment of self-reported sleep genes in Go Biological Process pathways

**Supplementary Table S19.** Edges (gene overlaps) between enrichment of the number of unique sleep medications genes in Go Biological Process pathways

**Supplementary Table S20.** Aggregated gene expression trajectory across developmental stages by sleep phenotype measurement categories.

## List of supplementary figures

**Supplementary Figure 1.** Gene discovery using gene-based (diamond), single rare variant (circle), single common variant (triangle) and meta-analyzed (square) associations for sleep phenotypes.

**Supplementary Figure 2.** Gene network enrichment in GO biological process pathways based on sleep genes. Each node represents an enriched GO term. Related GO terms are connected by a line, whose thickness reflects percent of overlapping genes. The size of the node corresponds to the number of genes, and the color of the node corresponds to the significance based on p values.

