## Supplementary Information for "Rare Coding Variants Reveal Distinct Genetic Architectures Across Multidimensional Sleep Phenotypes"

**Supplementary Figure 1.** Gene discovery using gene-based (diamond), single rare variant (circle), single common variant (triangle) and meta-analyzed (square) associations for sleep phenotypes.

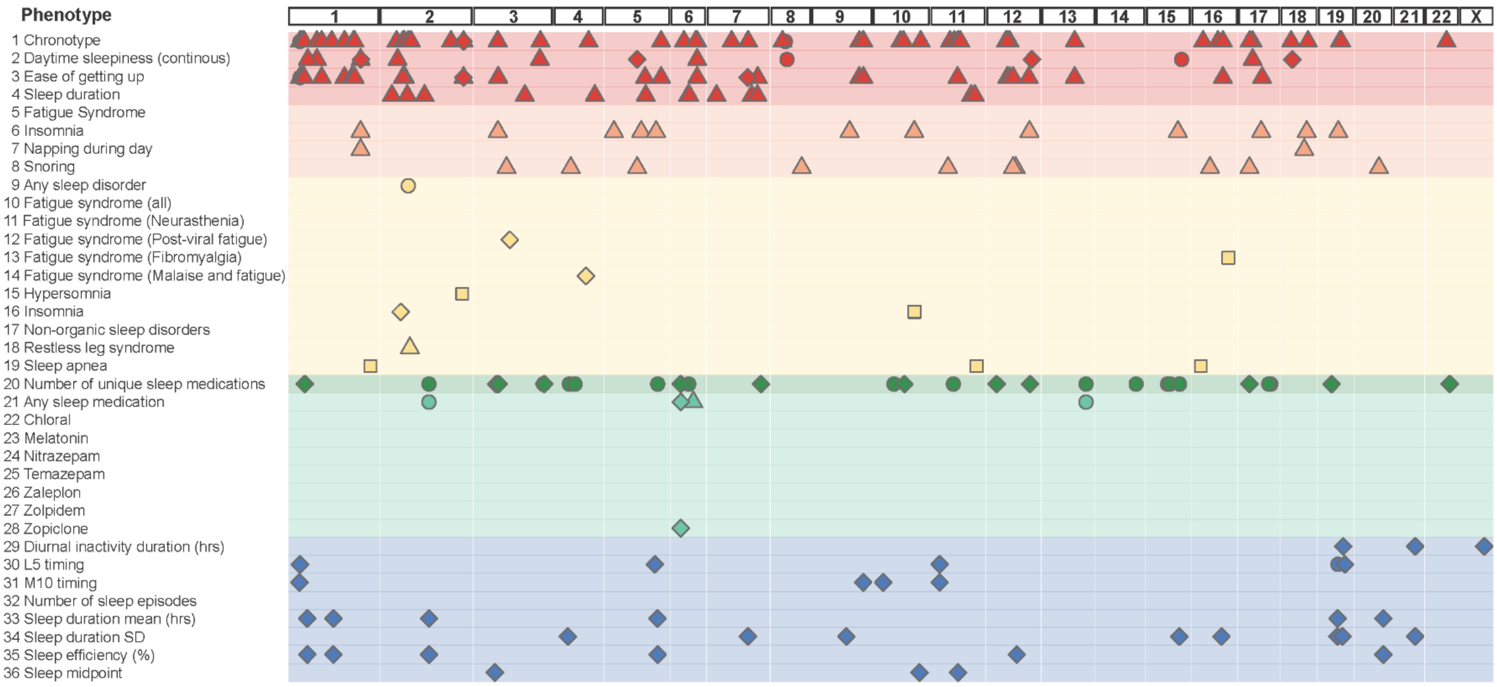

**Supplementary Figure 2.** Gene network enrichment in GO biological process pathways based on sleep genes. Each node represents an enriched GO term. Related GO terms are connected by a line, whose thickness reflects percent of overlapping genes. The size of the node corresponds to the number of genes, and the color of the node corresponds to the significance based on p values.

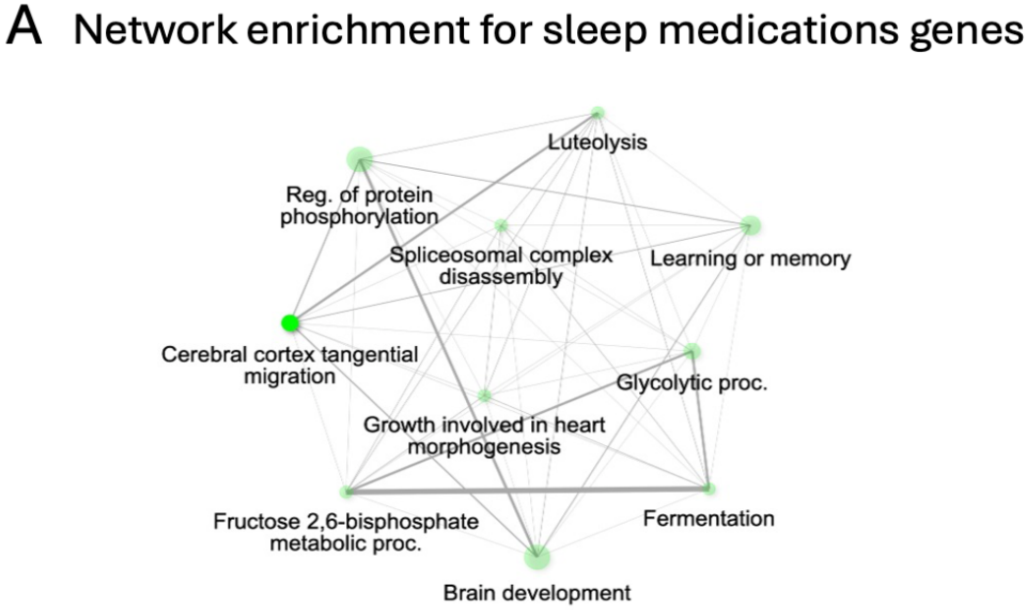


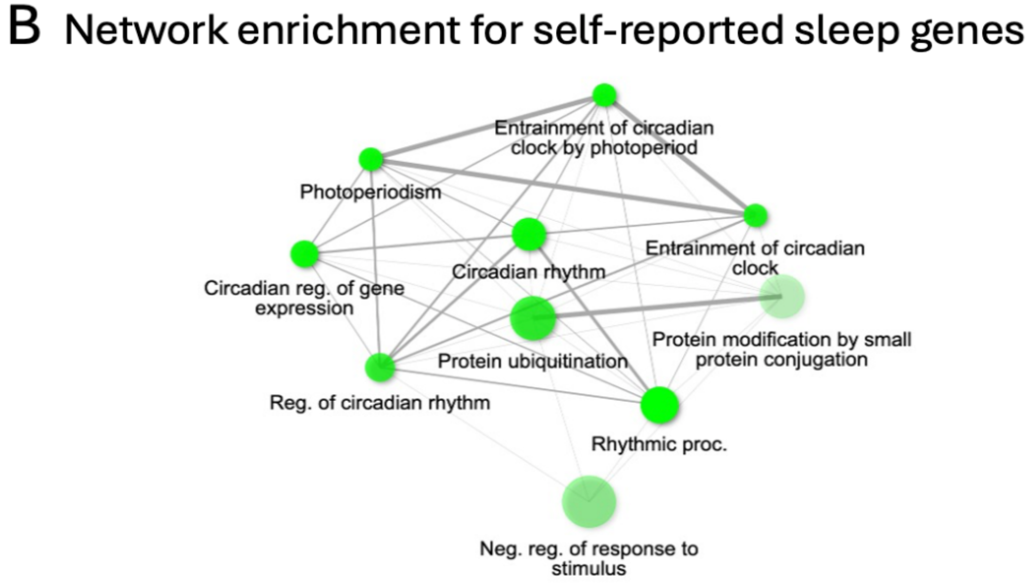
